# Effectiveness of Sotrovimab in Preventing COVID-19-related Hospitalizations or Deaths Among U.S. Veterans

**DOI:** 10.1101/2022.12.30.22284063

**Authors:** Yinong Young-Xu, Caroline Korves, Gabrielle Zwain, Sacha Satram, Myriam Drysdale, Carolina Reyes, Mindy M. Cheng, Lauren Epstein, Vincent C. Marconi, Adit Ginde

**Affiliations:** White River Junction Veterans Affairs Medical Center, White River Junction, VT; Geisel School of Medicine at Dartmouth, Hanover, NH; Vir Biotechnology, San Francisco, California, USA; GSK, Brentford, Middlesex, UK; Atlanta Veterans Affairs Medical Center, Decatur, GA; Division of Infectious Diseases, Emory University School of Medicine, Atlanta, GA; Department of Global Health, Rollins School of Public Health, Emory University, Atlanta, GA; Department of Emergency Medicine, University of Colorado School of Medicine, Aurora. CO

**Keywords:** COVID-19, SARS-CoV-2, monoclonal antibodies, prevention, real-world data

## Abstract

**Background:** Data on effectiveness of sotrovimab preventing COVID-19-related hospitalization or mortality, particularly after the emergence of the Omicron variant, are limited.

**Method:** Determine the real-world clinical effectiveness of sotrovimab for prevention of 30-day COVID-19 related hospitalization or mortality using a retrospective cohort within the U.S. Department of Veterans Affairs (VA) healthcare system.

Veterans aged ≥18 years, diagnosed with COVID-19 between December 1, 2021, and April 4, 2022, were included. Sotrovimab recipients (n=2,816) were exactly matched to untreated controls (n=11,250) on date of diagnosis, vaccination status, and region.

The primary outcome was COVID-19-related hospitalization or all-cause mortality within 30 days from diagnosis. Cox proportional hazards modeling estimated the hazard ratios (HR) and 95% Confidence Interval (CI) for the association between receipt of sotrovimab and outcomes.

**Results:** During BA.1 dominance, compared to matched controls, sotrovimab-treated patients had a 70% lower risk hospitalization within 30 days or mortality (HR 0.30; 95%CI, 0.23-0.40), a 66% lower risk of 30-day hospitalization (HR 0.34; 95%CI, 0.25-0.46), and a 77% lower risk of 30-day all-cause mortality (HR 0.23; 95%CI, 0.14-0.38). During BA.2 dominance sotrovimab-treated patients had a 71% (HR .29; 95%CI, 0.08-0.98) lower risk of 30-day COVID-19-related-hospitalization, emergency, or urgent care. Limitations include confounding by indication.

**Conclusions:** Using national real-world data from high risk and predominantly vaccinated Veterans, administration of sotrovimab, compared with no treatment, was associated with reduced risk of 30-day COVID-19-related hospitalization or all-cause mortality during the Omicron BA.1 period and reduced risk of progression to severe COVID-19 during the BA.2 dominant period.

**Summary:** Examination of national real-world evidence demonstrates sotrovimab is effective in preventing at risk positive COVID-19 cases from progressing to severe SARS-CoV-2 infections compared to matched untreated cases during Delta and early Omicron variant waves in the U.S. Veteran population.

## INTRODUCTION

Throughout the coronavirus disease 2019 (COVID-19) pandemic, Centers for Disease Control and Prevention (CDC) estimate that 96 million severe infections, 5 million hospitalizations, [1] and 1 million deaths were attributable to severe acute respiratory syndrome coronavirus 2 (SARS-CoV-2) in patients diagnosed with COVID-19 across the U.S. [2] From 2021 through 2022, SARS-CoV-2 variant tracking data showed the rise of the Delta variants during 2021 followed by Omicron variant dominance (BA.1, BA.2 and BA.2.12.1, BA 4/5) in 2022.

Sotrovimab is a recombinant human monoclonal antibody targeted against SARS-CoV-2, which received an Emergency Use Authorization (EUA) on May 26, 2021 for treatment of mild to moderate COVID-19 in adults and pediatric patients (12 years of age and older weighing at least 40 kg) at high-risk for progression to severe COVID-19 [3]. On March 25, 2022, due to increasing prevalence of the SARS-CoV-2 Omicron BA.2 sub-variant and concern for resistance against sotrovimab (Appendix Table 1) [4], sotrovimab was de-authorized by the FDA in any U.S. region with Omicron BA.2 prevalence >50%. Sotrovimab was de-authorized for the entire country on April 5, 2022.

Among outpatients with mild to moderate COVID-19 at risk of disease progression, a single 500mg intravenous dose of sotrovimab, compared with placebo, was associated with significant reduction in the risk of a composite end point of 29-day all-cause hospitalization or death (COMET-ICE) [**Error! Bookmark not defined**.]. These findings supported sotrovimab as a treatment option for high-risk outpatients with mild to moderate COVID-19 caused by SARS-CoV-2 variants circulating through March 2021: Alpha, Beta, Gamma, Delta, and Lambda [5,6]. Some regional retrospective analysis has been done; however, examination of national real-world evidence during Delta and Omicron predominant periods is needed to determine ongoing effectiveness of sotrovimab treatment in high-risk COVID-19 patients [7,8].

Our objective was to assess the effectiveness of sotrovimab for reduction of COVID-19 related hospitalization, or all-cause mortality within 30 days of treatment during the period of Omicron dominance using electronic health data from the U.S. Department of VA, the largest integrated health care system in the U.S.

## METHODS

### Study Setting and Data Sources

VA provides care to nearly 9 million Veterans at 171 medical centers and 1,113 outpatient clinics across the U.S. We analyzed electronic health records (EHR) using the VA Corporate Data Warehouse, which contains patient-level information on all clinical encounters in VA medical facilities, including treatments, prescriptions, vaccinations, laboratory results, healthcare utilization, and vital status [9,10]. We identified sotrovimab use through the VA Pharmacy Benefits Management EUA prescription dashboard, which captures and links records of recipients, date, and dosage of sotrovimab administered in medical facilities across the VA [11]. Sotrovimab was available for administration at the VA from December 2021 to April 2022; first dose administered on December 1^st^, 2021, and last dose on April 4^th^, 2022.

Before data collection, this study was approved by the institutional review board of the VA Medical Center in White River Junction, Vermont and was granted a waiver of informed consent because the study was deemed minimal risk and consent impractical to acquire. This study followed the Strengthening the Reporting of Observational Studies in Epidemiology (STROBE) reporting guideline.

### Study Population and Outcomes

We included Veterans who were ≥18 years, diagnosed with COVID-19 (SARS-CoV-2 detected via antigen or PCR testing) between December 1^st^, 2021 through April 4, 2022 and received Veterans Health Administration (VHA) benefits for at least 2 years prior to diagnosis. Because our study focused on the effect of sotrovimab versus no treatment, we included positive lab test and documented home testing of Veterans with SARS-CoV-2 infection. We identified patients not requiring hospitalization or new supplemental oxygen, yet at high-risk of disease progression, in line with sotrovimab EUA eligibility criteria, using diagnosis codes during the 2 years prior to their SARS-CoV-2 infection diagnosis date. Among eligible individuals we identified patients who received sotrovimab as treated. Controls were selected in regions where sotrovimab was approved for use among eligible patients who did not receive any early antiviral treatment (oral antivirals or monoclonal antibodies). Index date was first of the positive SARS-CoV-2 lab test date or documented home test. If either is missing, for sotrovimab recipients, index date was their diagnosis date for COVID-19 (ICD 10 code U07.1).

We included baseline characteristics (e.g., demographics, significant comorbidities, and healthcare utilization) documented within 2 years prior to the index date. We used the VA-assigned priority group for healthcare to serve as a surrogate measure for socioeconomic status [12]. Information regarding comorbidities was extracted from diagnostic codes recorded in VA electronic data for healthcare encounters; significant comorbidities were defined according to an adaptation of Deyo-Charlson comorbidity index (DCCI) [13].

We divided the study period into 3 predominant variant periods: Delta dominance (December 1^st^ to December 17^th^, 2021), BA.1 dominance (from December 18^th^, 2021, to March 15^th^, 2022), and BA.2 dominance (starting March 16th, 2022, lasting until the end of the study period, May 4^th^, 2022) (Appendix Figure 1). Our main analysis focuses on the BA.1 period, and the primary outcome was the composite of 1) COVID-19 hospitalization, defined as having both an admission and discharge diagnosis for COVID-19 from a hospital within 30 days of the index (e.g., positive SARS-CoV-2 test) date or 2) all-cause mortality, defined as having a date of death during the follow-up, also within 30 days of the index date. The time to event was the time from index date to first occurrence of the outcome of interest (30-day hospitalization or mortality). We studied the effectiveness of sotrovimab against the composite outcome stratified by age and high-risk groups - 65 or older, patients who are immunocompromised or with renal disease, chronic obstructive pulmonary disease (COPD), or cardiovascular disease. We used the outcome of clinical visits with urinary tract infection (UTI) as a falsification test or negative control [14]. We chose UTI as a negative control because there is no causal mechanism for an association between sotrovimab and UTI, but some of the same biases that exist for association between sotrovimab and UTIs could be similar to those between sotrovimab and COVID-19 outcomes [15].

### Statistical Analysis

#### Matching

First, among treated and eligible controls, we further restricted to those who met at least one criterion for sotrovimab use under the EUA [16]. We then used exact matching to account for observable baseline differences between patients who received sotrovimab and untreated controls. Exact matching was implemented for the following variables: SARS-CoV-2 diagnosis date for treated and controls within one week, vaccination status (unvaccinated, vaccinated with a primary series (2 mRNA or a single Janssen) or primary series plus booster), and Health & Human Services (HHS) region because the FDA de-authorized sotrovimab dissemination by HHS region following the rise of Omicron BA.2 variant (Appendix Table 1). We exact matched on certain underlying conditions: chronic kidney disease or renal disease, immunosuppressive disease, or immunosuppressive treatment, chronic obstructive pulmonary disease (most prevalent chronic lung disease in VA) or asthma, and cardiovascular disease or hypertension.

We included all the additional variables (**Table 1**) that were not included in the exact matching as covariates in our survival analyses. Covariates were measured before the initiation of sotrovimab to avoid adjustment for potential mediators. Indicator variables were generated to capture missing or unknown values for any of the matching criteria to retain patients in the study. To assess the robustness of the matching, we calculated standardized mean difference (SMD) and a difference of 10 or greater was used to identify imbalance between treated and controls.

**Table 1.**
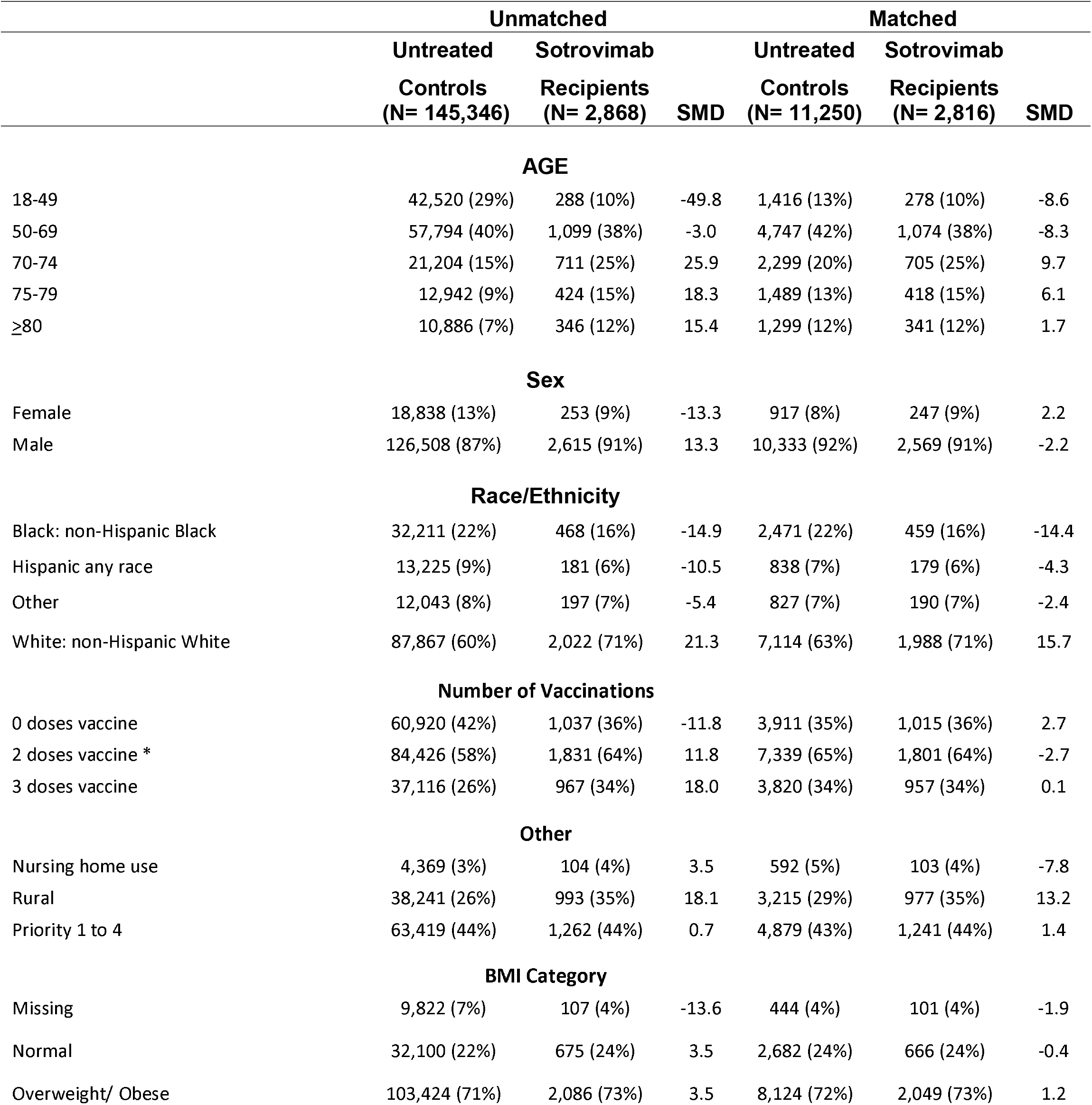
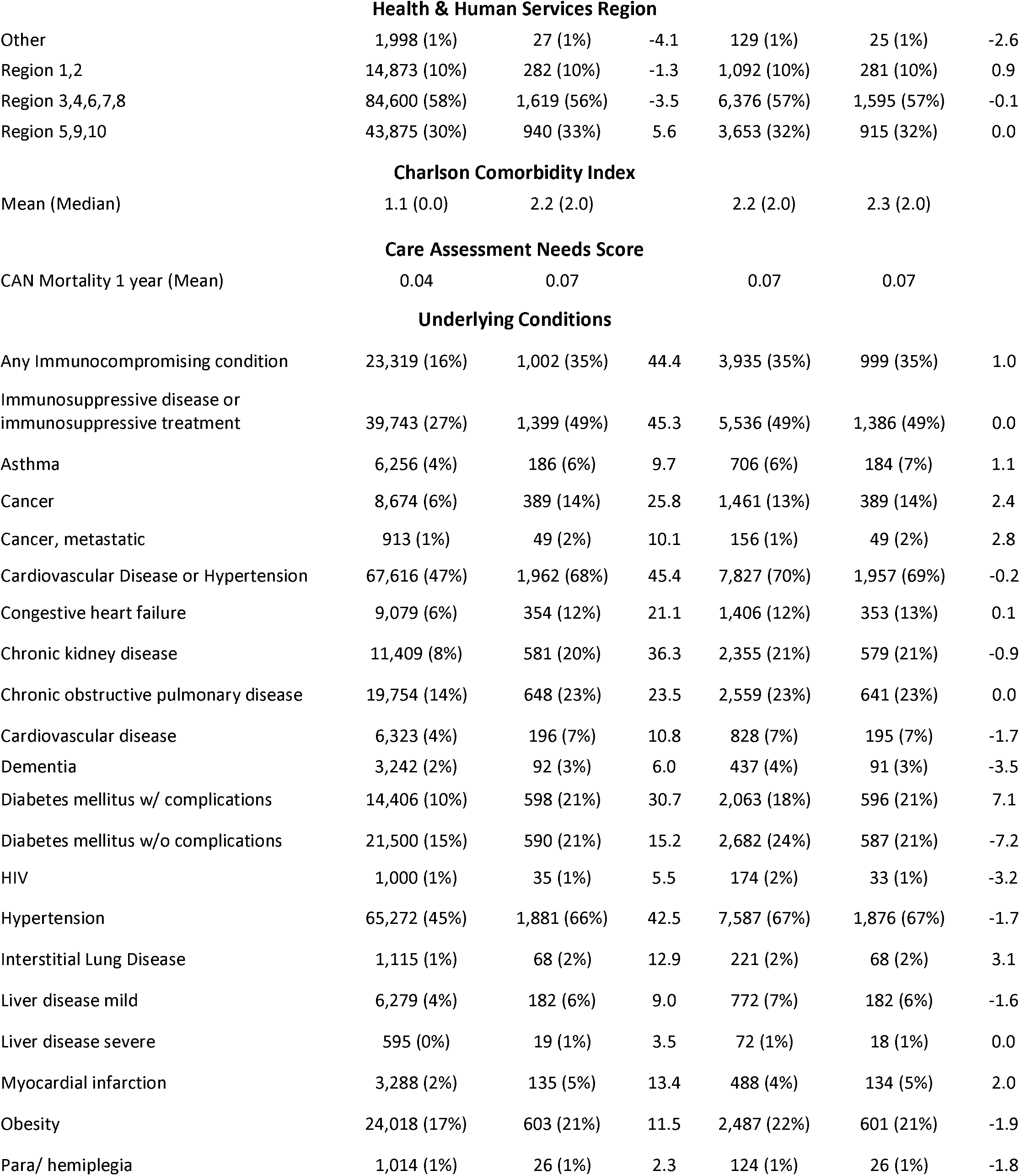
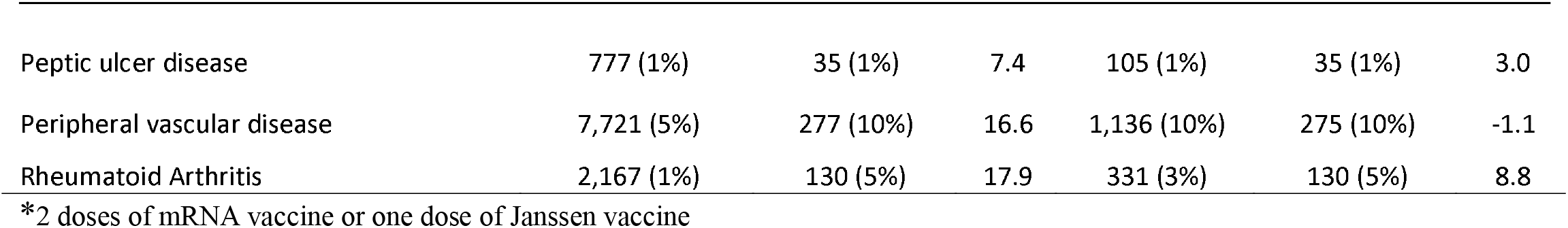
Baseline Characteristics

#### Censoring

We used Cox proportional hazards regression model to compare patients who received sotrovimab and their matched untreated controls. Since outcomes were assessed for the 30-day period after index date, patients were censored on the earliest of: May 4^th^, 2022 (earliest date to allow 30-day post-index observation period for the last sotrovimab given in VA), disenrollment, death, or 30 days post-index.

#### Secondary Analyses

The FDA began to de-authorize sotrovimab on March 25^th^, 2022 [Appendix Table 1], shortly after BA.2 became the predominant SARS-CoV-2 variant in the U.S. Consequently, during this period of BA.2 predominance, we observed few sotrovimab administrations and fewer outcomes through April 2022. Similarly, we have a short Delta period (17 days) starting when sotrovimab became available at VA and ending when Omicron BA.1 became predominant, resulting in a small number of primary outcomes. Since these periods were both short, we included COVID-19 related-emergency room (ED) or urgent care (UC) visits as a surrogate for persistent or severe COVID-19 for our effectiveness estimate of sotrovimab against either Delta or Omicron BA.2 SARS-CoV-2 variants. The time to event was the index date to the first occurrence of the outcome of interest (30-day hospitalization or ED visits or UC visits).

Analyses were performed with Stata 17 software (StataCorp), and SAS software, version 8.2 (SAS Institute).

## RESULTS

### Study Population

We identified 2,868 sotrovimab recipients and 145,346 controls. Representative of VA population, we found among sotrovimab recipients most were non-Hispanic White (71%) males (91%) over the age of 50 (90%) living in urban areas [17] (65%), the majority (64%) of whom had received 2 doses of mRNA vaccines or 1 dose of Ad26.COV2 (Janssen). Common comorbidities among sotrovimab recipients included hypertension (1,876 (67%)), and any immunocompromising condition or immunosuppressive treatment (1,386 (49%)). After exact matching, 2,816 sotrovimab recipients and 11,250 control patients were balanced across baseline characteristics (**Table 1**).

### Matched Analysis

#### Main analysis/BA.1 period

Sotrovimab recipients had a lower incidence of the composite COVID-19 outcome 92/2557 [3.6%] vs untreated cases 735/10297 [7.1%]; and a 70% lower risk (HR 0.30; 95%CI, 0.23-0.40) of the composite 30-day hospitalization or all-cause mortality outcome compared to controls. Sotrovimab was associated with a similar risk reduction in the composite outcome among those 65 years of age or older (HR 0.33; 95% CI, 0.24-0.45) as well as among those immunocompromised or those who have renal disease (HR 0.30; 95% CI,0.20-0.45).

Overall, sotrovimab recipients also had a 66% lower risk of COVID-19 related 30-day hospitalization (HR 0.34; 95% CI 0.25-0.46) and a 77% lower risk of 30-day all-cause mortality (HR 0.23; 95% CI, 0.14-0.38). (**Table 2**)

**Table 2.**
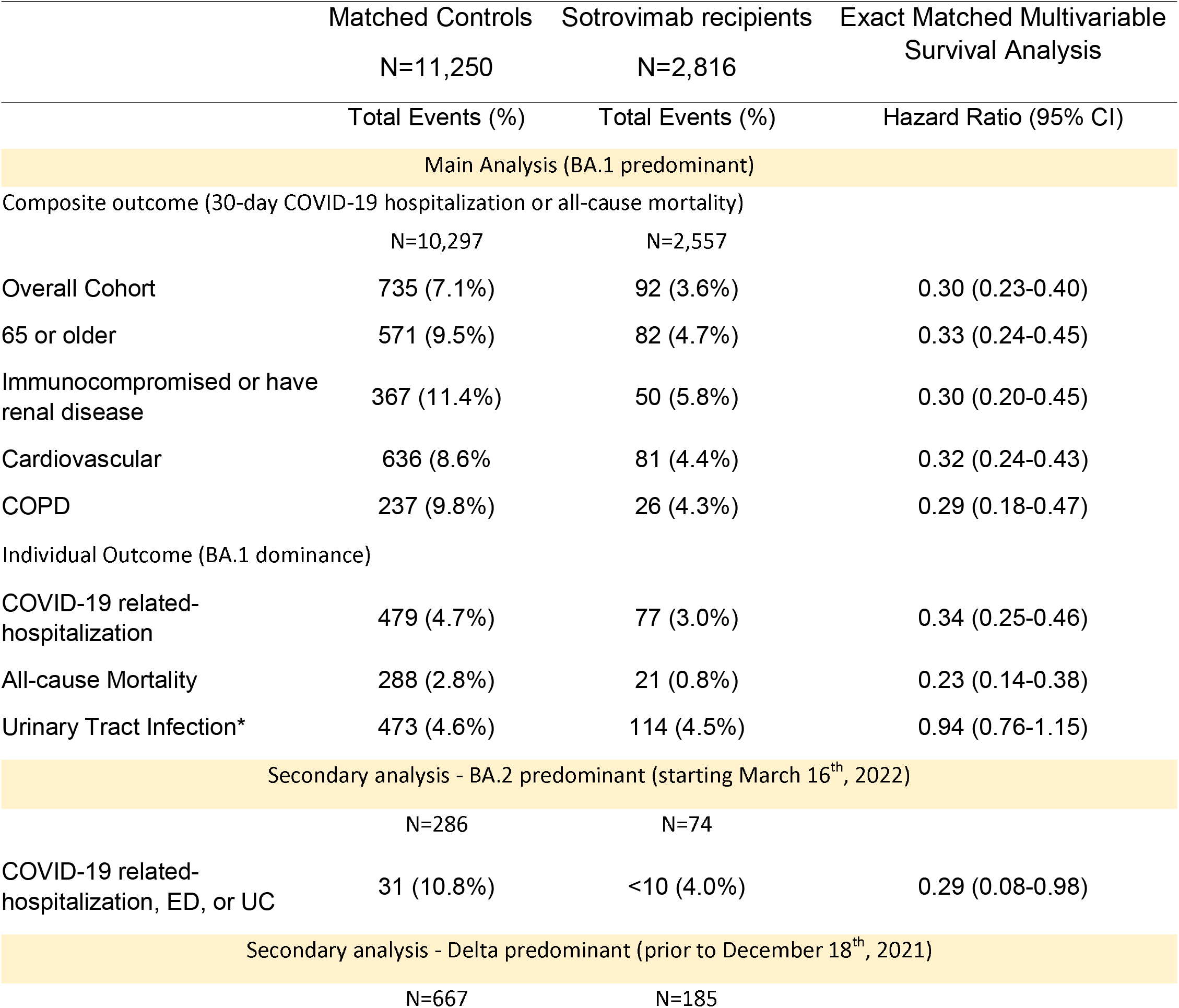
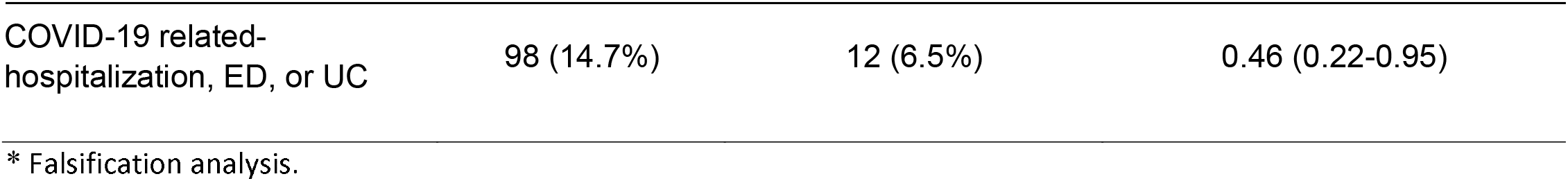
Relative Effectiveness of Sotrovimab versus Untreated Controls using Adjusted Analysis

|  | Matched Controls<br>N=11,250 | Sotrovimab recipients<br>N=2,816 | Exact Matched Multivariable<br>Survival Analysis |
| --- | --- | --- | --- |
|  | Total Events (%) | Total Events (%) | Hazard Ratio (95% CI) |
| Main Analysis (BA.1 predominant) |  |  |  |
| Composite outcome (30-day COVID-19 hospitalization or all-cause mortality) |  |  |  |
|  | N=10,297 | N=2,557 |  |
| Overall Cohort | 735 (7.1%) | 92 (3.6%) | 0.30 (0.23-0.40) |
| 65 or older | 571 (9.5%) | 82 (4.7%) | 0.33 (0.24-0.45) |
| Immunocompromised or have renal disease | 367 (11.4%) | 50 (5.8%) | 0.30 (0.20-0.45) |
| Cardiovascular | 636 (8.6%) | 81 (4.4%) | 0.32 (0.24-0.43) |
| COPD | 237 (9.8%) | 26 (4.3%) | 0.29 (0.18-0.47) |
| Individual Outcome (BA.1 dominance) |  |  |  |
| COVID-19 related-hospitalization | 479 (4.7%) | 77 (3.0%) | 0.34 (0.25-0.46) |
| All-cause Mortality | 288 (2.8%) | 21 (0.8%) | 0.23 (0.14-0.38) |
| Urinary Tract Infection* | 473 (4.6%) | 114 (4.5%) | 0.94 (0.76-1.15) |
| Secondary analysis - BA.2 predominant (starting March 16<sup>th</sup>, 2022) |  |  |  |
|  | N=286 | N=74 |  |
| COVID-19 related-hospitalization, ED, or UC | 31 (10.8%) | <10 (4.0%) | 0.29 (0.08-0.98) |
| Secondary analysis - Delta predominant (prior to December 18<sup>th</sup>, 2021) |  |  |  |
|  | N=667 | N=185 |  |
| COVID-19 related-hospitalization, ED, or UC | 98 (14.7%) | 12 (6.5%) | 0.46 (0.22-0.95) |

\* Falsification analysis.

#### Secondary Analyses/Delta and BA.2 periods

Sotrovimab recipients during the Delta dominance period (December 1^st^ through December 17^th^, 2021) had few outcomes and a 54% lower risk of COVID-19 related-hospitalization, emergency department, or urgent care visits (HR 0.46; 95% CI, 0.22-0.95). Lastly, we examined the impact of sotrovimab during the period of BA.2 and BA.2.12.1 dominance, based on CDC variants tracking. From March 16 through April 4, 2022, 74 patients received sotrovimab. Compared to matched controls, sotrovimab recipients had a 71% lower risk of COVID-19 related-hospitalization, emergency department, or urgent care visits (HR 0.29; 95% CI, 0.08-0.98) during the period of BA.2 and BA.2.12.1 dominance. (**Table 2)** Sequencing data are not available for these patients.

### Falsification Analysis

Six hundred ninety-seven UTI visits were observed during the follow-up period. Matched analysis demonstrated a similar effectiveness of sotrovimab versus control against UTI (HR 0.94; 95% CI, 0.76-1.15) (**Table 2**). This lack of association between sotrovimab and UTI confirmed our assumption that the protective effects of sotrovimab administration were unlikely due to bias or other major methodological flaws.

## DISCUSSION

In this retrospective cohort study using data from patients across the VA healthcare system in the U.S., administration of sotrovimab was associated with a significant reduction in the risk of COVID-19 hospitalization and all-cause mortality compared with untreated controls in exact matched analysis. Findings were consistent among immunocompromised, patients with renal diseases, and those who were 65 years or older. Furthermore, we did not see evidence of reduced effectiveness during the rise of SARS-CoV-2 BA.2 variant, though limited by sample size due to sotrovimab no longer being authorized for use in the U.S. after BA.2 was predominant. This study provides important insights regarding the effectiveness of sotrovimab across the VA health care system and our results are consistent with real-world studies of sotrovimab effectiveness from other U.S. populations.

Aggarwal and colleagues examined effectiveness of sotrovimab among high-risk outpatients diagnosed with COVID-19 in Colorado during October 1, 2021 – December 11, 2021 when Delta was the predominant circulating variant and found a 63% lower odds of 28-day all-cause hospitalization and 89% lower odds of 28-day all-cause mortality[7]. A subsequent study assessed sotrovimab effectiveness among high-risk COVID-19 patients between December 26, 2021 – March 10, 2022 and found non-significant lower odds of 28-day all-cause hospitalization or mortality during the Omicron BA.1 and BA.2 waves due to the smaller sample size: “28-day hospitalization (2.5% versus 3.2%; adjusted OR 0.82, 95% CI 0.55, 1.19) or mortality (0.1% versus 0.2%; adjusted OR 0.62, 95% CI 0.07, 2.78)” [8 Finally, a large representative U.S. sample of high-risk COVID-19 patients diagnosed between September 1, 2021 to April 30, 2022 in the FAIR Health National Private Insurance Claims database found that patients receiving sotrovimab had a 55% lower risk of 30-day hospitalization or mortality (RR 0.45, 95% CI, 0.41 - 0.49) and an 85% lower risk of 30-day mortality (RR 0.15, 95% CI, 0.08 - 0.29) [18]. Like Aggarwal et al, Cheng et al found reduced, non-significant sotrovimab effectiveness during BA.2 predominance – RR 0.32, 95% CI, 0.04 - 2.38 – in April 2022, with 68 doses of sotrovimab dispensed in an eligible population over 117 thousand.

With the emergence of the Omicron BA.2 subvariant in early 2022, the US Food and Drug Administration (FDA) found a decrease in microneutralisation titre EC90 of 25–48-fold relative to ancestral SARS-CoV-2[4]. On April 5, 2022, the FDA withdrew approval of sotrovimab when in vitro neutralization results and conservative assumptions were used in pharmacokinetic modelling suggesting that the authorized dose was unlikely to be effective against BA.2 [4]. This decision is not supported by real-world data. Piccicacco et al., found that “Patients receiving sotrovimab were also less likely to be hospitalized or visit the ED (8% versus 23.3%; OR=0.28, 95% CI, 0.11–0.71).” [19] We found almost the same effectiveness of 0.29; 95% CI,0.08-0.98 for COVID-19 related-hospitalization, ED, or UC, although we had a smaller sample size (sotrovimab n=74), thus wider interval. Zheng et al., studied sotrovimab effectiveness among a large cohort with similar demographics and again among patients on kidney replacement therapy covering BA.2 predominant period in the UK and found sotrovimab to be effective throughout BA.1, BA. 2 and BA.4/5 predominance [20].

In the case of sotrovimab, combined evidence from our study and others using real-world clinical effectiveness data supports its continued use against circulating Omicron variants, including BA.4 and BA.5. The ongoing evolution of SARS-CoV-2 variants and continued global transmission has resulted in a situation where new variants can replace one another within weeks. This is a challenging environment for regulatory agencies as clinical data research takes time while in-vitro results might not be applicable outside a petri dish. Examination of real-world patient data will avoid scenarios wherein effective monoclonals are available but not offered to clinically vulnerable patients.

Our study has several notable strengths. We analyzed 1,669 patient-years of observation, making our study one of the largest conducted using electronic medical records to assess sotrovimab effectiveness while it was being utilized to combat a concurrent surge of the pandemic. The large sample allowed us to adjust for more potential confounding variables. Previous studies have shown that EHR data are more likely than insurance claims data to be complete in capturing medical conditions and have a lower risk of up-coding [21,22]. Nevertheless, conventional analytical strategies such as stratification, matching (with or without propensity score), and multivariate regression analysis cannot adequately adjust for unmeasured confounders [23,24,25]. Because more than 90% of the time VA provided sotrovimab to patients within 1 to 2 days of their infection, matching those treated and untreated on the day of their infection ensured similar lengths of follow-up between the recipients and their matched controls thus reduced immortal bias.

### Limitations

First, VA data include only health care encounters that occur in VA medical centers, so we could have missed hospitalizations that occurred outside the VA (ascertainment bias); assuming missed events were as likely among those treated and controls this non-differential misclassification would have biased our results toward the null. Second, VA has a unique population (mostly male, older), and our results may not be generalizable to a larger population of patients not treated at VA (selection bias) [26]. Third, the 10th revision of the International Statistical Classification of Diseases and Related Health Problems (ICD-10) codes from claims data have been shown to inadequately capture comorbidity and functional status [27]. Fourth, we did not have sequencing data for this study to determine the variants of the SARS-CoV-2 infections included in this analysis. While our study reported COVID-19 treatments given following sotrovimab administration, the impact of these treatments on the estimated association of sotrovimab and severe COVID-19 outcomes was not assessed, and it is possible those differences may have affected differences in hospitalization and mortality rates between cohorts. We matched treated and control patients in regions where sotrovimab was given to ensure comparability between groups for this study, but further research on drivers of prescribing differences is needed as to why patients may not realize the potential benefit and eligibility for this and other COVID-19 treatments [28].

Finally, although it is likely half of the infections could be due to BA.2 during the last month of our study period based on CDC variants tracking data, we plan to use sequencing data in our next study to better assess sotrovimab against BA.2.

## Conclusion

Using national data from Veterans, we found administration of sotrovimab was associated with lower risks of 30-day COVID-19 related hospitalization and all-cause mortality, compared with controls, during the period of Omicron BA.1 dominance. Our real-world results also suggest that sotrovimab administration may protect vulnerable patients from severe COVID-19 during BA.2 dominance. Ongoing real-world data will help to understand the clinical effectiveness of sotrovimab over time and against emerging variants. These results indicate the monoclonal antibody therapy is an effective strategy for treatment of COVID-19 for certain patient populations with susceptible dominant SARS-CoV-2 strains.

The contents of this article do not represent the views of the U.S. Department of Veterans Affairs or the U.S. Government. The views and opinions expressed in this article are those of the authors and do not necessarily reflect the official policy or position of the Food and Drug Administration, as well as any other agency of the U.S. Government. Assumptions made within and interpretations from the analysis do not necessarily reflect the position of any U.S. Government entity.

## Data Availability

Requests for access to the de-identified study data can be sent via e-mail to. No supporting documents will be made available. The data will be made available to qualified scientific researchers for specific purposes outlined in a proposal after the researcher enters into a standard data sharing agreement and the proposal is approved. Researchers must commit to transparency in publication.

## Author, Article and Disclosure Information

Affiliations

White River Junction Veterans Affairs Medical Center, White River Junction, VT (Y.Y.X., G.Z., J.S., C.K.)

Geisel School of Medicine at Dartmouth, Hanover, NH (Y.Y.X.)

Atlanta Veterans Affairs Medical Center, Decatur, GA (L.E., V.C.M.)

Division of Infectious Diseases, Emory University School of Medicine, Atlanta, GA (V.C.M.)

Department of Global Health, Rollins School of Public Health, Emory University, Atlanta, GA (V.C.M.)

U.S. Department of Veteran’s Affairs, Office of Research and Development, Washington DC (V.D.)

U.S. Department of Veterans Affairs, PBM, Center for Medication Safety, Hines, IL (F.C.)

U.S. Department of Veterans Affairs, VA SHIELD, Veterans Affairs Northeast Ohio Healthcare System, Cleveland, Ohio, USA (R.A.B.)

Case Western Reserve University, Cleveland, Ohio, USA (R.B.)

Department of Emergency Medicine, University of Colorado School of Medicine, Aurora. CO (A.A.G.)

## Acknowledgment

Medical writing assistance was provided by Tara Krause, BA of Clinical Epidemiology Program at the White River Junction VA Medical Center, Vermont, USA.

## Data Sharing Statement

Requests for access to the deidentified study data can be sent via e-mail to. No supporting documents will be made available. The data will be made available to qualified scientific researchers for specific purposes outlined in a proposal after the researcher enters into a standard data sharing agreement and the proposal is approved. Researchers must commit to transparency in publication.

## Author Contributions

Conception and design: Y. Young-Xu, C. Korves, G. Zwain, M. Drysdale, S. Satram, C. Reyes, M.M.Cheng, L. Epstein, V.C. Marconi, A.A. Ginde

Analysis and interpretation of the data: Y. Young-Xu, C. Korves, G. Zwain, M. Drysdale, S. Satram, C. Reyes, M.M.Cheng, L. Epstein, V.C. Marconi, A.A. Ginde

Drafting of the article: Y. Young-Xu, C. Korves, G. Zwain

Critical revision for important intellectual content: Y. Young-Xu, C. Korves, G. Zwain, M. Drysdale, S. Satram, C. Reyes, M.M.Cheng, L. Epstein, V.C. Marconi, A.A. Ginde

Final approval of the article: Y. Young-Xu, C. Korves, G. Zwain, M. Drysdale, S. Satram, C. Reyes, M.M.Cheng, L. Epstein, V.C. Marconi, A.A. Ginde.

Provision of study materials or patients: Y. Young-Xu

Statistical expertise: Y. Young-Xu., C. Korves

Administrative, technical, or logistic support: Y. Young-Xu.

Collection and assembly of data: Y. Young-Xu, G. Zwain

---

**Figure 1.**
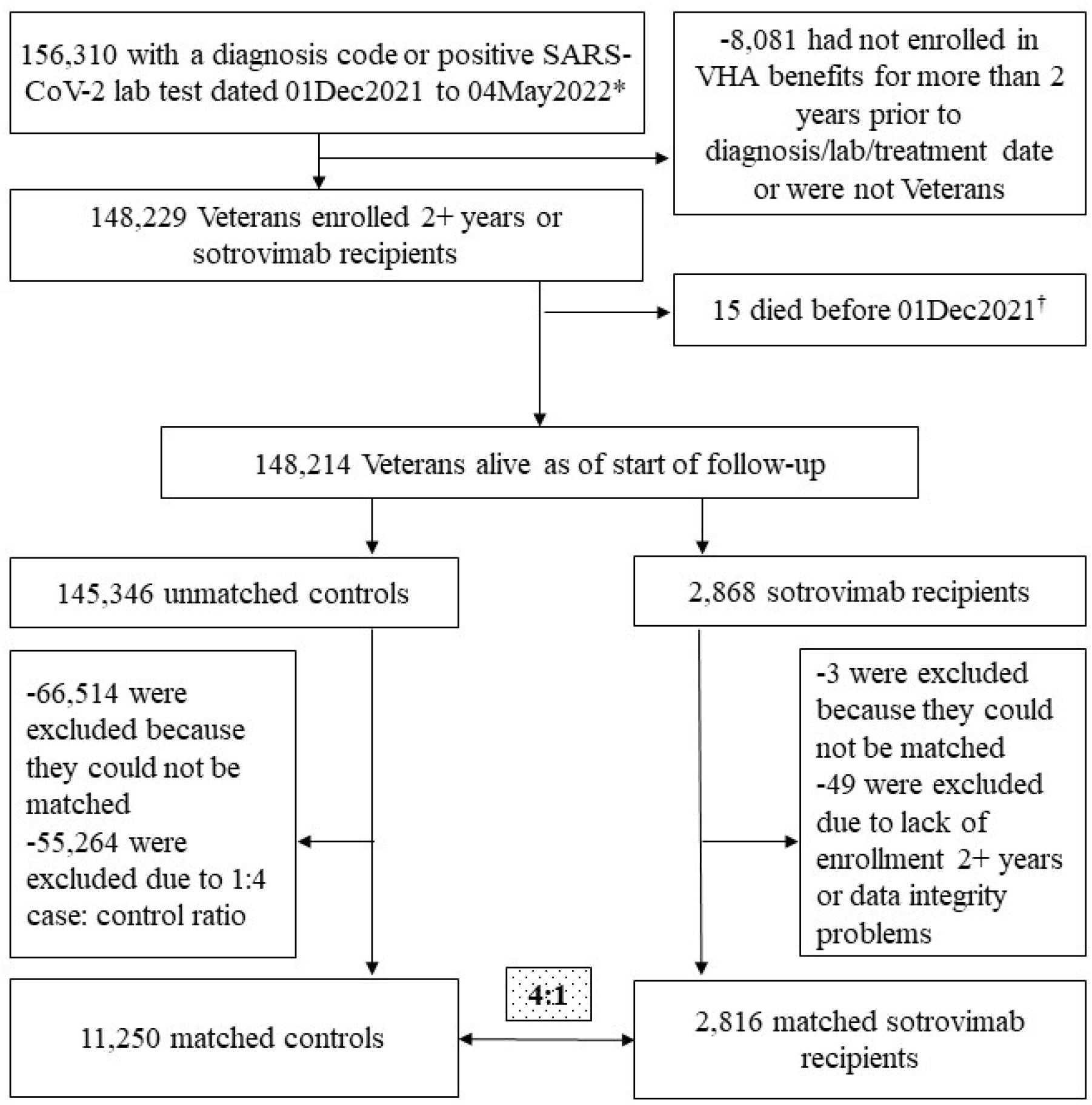
Attrition. *Includes those with a sotrovimab prescription and presumed positive lab test (47 patients without a visible lab or dx) Exact matching criteria: Had booster at index, control was alive as of the case’s index date, sotrovimab prescription date and control’s diagnosis/ positive lab date was within one week of each other. HHS category: (HHS regions 1 or 2, HHS regions 5, 9, or 10, HHS regions 3, 4, 6, 7, 8), Comorbid condition category: kidney (CKD, Renal), lung (COPD, Asthma), immunosuppressive prescription or diagnosis, cardiac diagnosis (CAD or HTN) Controls had to have one of the following conditions: CKD, Renal Disease, Diabetes, CAD, MI, CHF, Dyslipidemia, ILD, Sickle Cell Disease, Immunocompromising Condition, COPD, Asthma, Hypertension, CAD, Hematological Cancers) †One person had problems with the Date of Death (it was before treatment and before many other records (vitals, labs, etc.) This person was excluded due to data issues.

**Figure 2.**
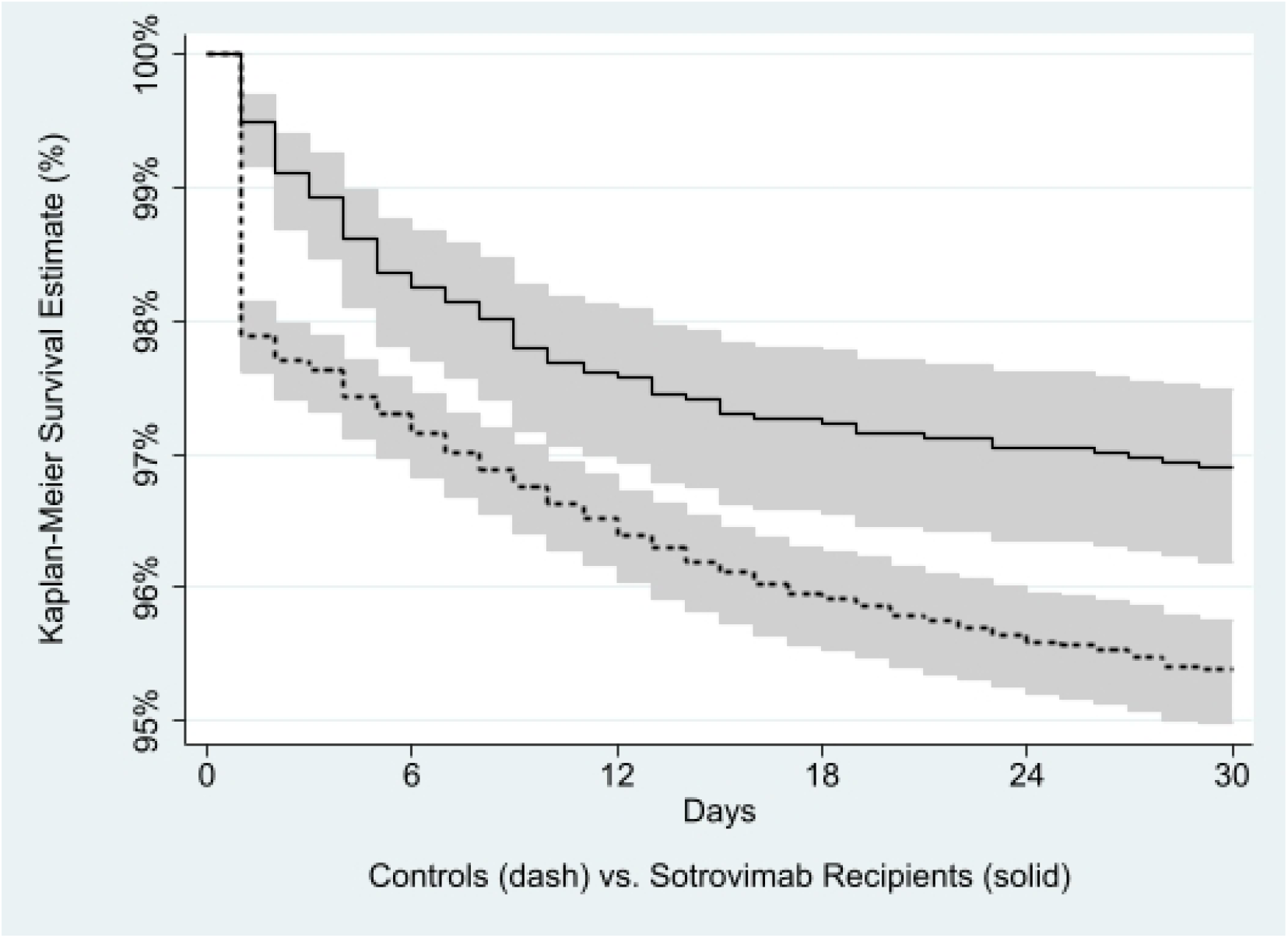
Kaplan-Meier curve with 95% confidence bands showing survival rate (%) of COVID-19-related hospitalizations or deaths in Veterans who received sotrovimab versus matched untreated controls.

## Appendix

**Table 1**. A Brief Timeline Related to Sotrovimab (starting with the most recent):

- May 12, 2022: FDA Authorizes Shelf-Life Extension for Sotrovimab From 12 to 18 Months
- April 5, 2022: Distribution of Sotrovimab Paused to All Department of Health & Human Services (HHS) Regions
- March 30, 2022: Distribution of Sotrovimab Paused to Certain States (HHS Region 5, Region 9, and Region 10)
- March 25, 2022: Distribution of Sotrovimab Paused to Certain States (HHS Region 1 and Region 2)
- December 17, 2021: The U.S. federal government prepared approximately 55,000 doses of sotrovimab for immediate allocation to jurisdictions. Jurisdictions saw products arrive as early as Tuesday, December 21, 2021.
- In late November 2021, the federal government paused shipment of the monoclonal antibody therapeutic sotrovimab to help ensure a more balanced portfolio of monoclonal antibody products and to allow more time to assess data regarding the effectiveness of sotrovimab against the Omicron variant.[28]
- May 26, 2021: FDA EUA for Sotrovimab

**Table 2**. High-Risk Conditions (reference 14, FACT SHEET FOR HEALTHCARE PROVIDERS

Older age (for example ≥65 years of age) • Obesity or being overweight (for example, adults with BMI >25 kg/m2, or if 12 to 17 years of age, have BMI ≥85th percentile for their age and gender based on CDC growth charts, https://www.cdc.gov/growthcharts/clinical_charts.htm) • Pregnancy • Chronic kidney disease • Diabetes 2 •Immunosuppressive disease or immunosuppressive treatment • Cardiovascular disease (including congenital heart disease) or hypertension • Chronic lung diseases (for example, chronic obstructive pulmonary disease, asthma [moderate-to-severe], interstitial lung disease, cystic fibrosis and pulmonary hypertension) • Sickle cell disease • Neurodevelopmental disorders (for example, cerebral palsy) or other conditions that confer medical complexity (for example, genetic or metabolic syndromes and severe congenital anomalies) • Having a medical-related technological dependence (for example, tracheostomy, gastrostomy, or positive pressure ventilation [not related to COVID-19])

**Figure 1.**
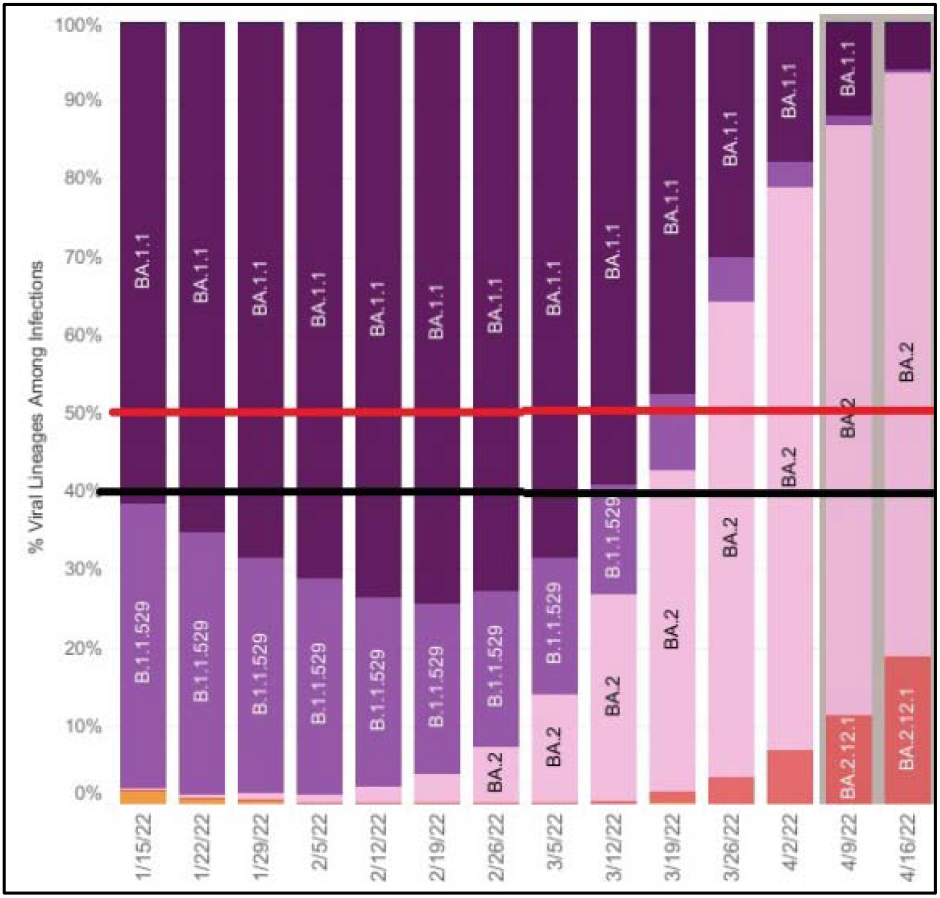
Identified study periods based on variant lineage emergence.

